# Case-control, diagnostic accuracy study of a non-sputum CD38-based TAM-TB test from a single milliliter of blood

**DOI:** 10.1101/2021.03.10.21253246

**Authors:** Hellen Hiza, Jerry Hella, Ainhoa Arbués, Beatrice Magani, Mohamed Sasamalo, Sebastien Gagneux, Klaus Reither, Damien Portevin

## Abstract

**Background:** CD4 T cell phenotyping-based blood assays have the potential to meet WHO target product profiles (TPP) of non-sputum-biomarker-based tests to diagnose tuberculosis (TB). Yet, substantial refinements are required to allow their implementation in clinical settings. This study assessed the real time performance of a simplified T cell activation marker (TAM)-TB assay to detect TB in adults from one millilitre of blood with a 24h turnaround time.

**Methods:** We recruited 479 GeneXpert^®^ positive cases and 108 symptomatic but GeneXpert^®^ negative controls from presumptive adult TB patients in the Temeke District of Dar-es-Salaam, Tanzania. TAM-TB assay accuracy was assessed by comparison with a composite reference standard comprising GeneXpert^®^ and solid culture. A single millilitre of fresh blood was processed to measure expression of CD38 or CD27 by CD4 T cells producing INF-γ and/or TNF-α in response to a synthetic peptide pool covering the sequences of *Mycobacterium tuberculosis* (*Mtb*) ESAT-6, CFP-10 and TB10.4 antigens on a 4-color FACSCalibur apparatus.

**Results:** Significantly superior to CD27 in accurately diagnosing TB, the CD38-based TAM-TB assay specificity reached 93.4% for a sensitivity of 82.2% with an area under the receiver operating characteristics curve of 0.87 (95% CI 0.84-0.91). The assay performance was not significantly affected by HIV status.

**Conclusions:** We successfully implemented TAM-TB immunoassay routine testing with a 24h turnaround time at district level in a resource limited setting. Starting from one millilitre of fresh blood and being not influenced by HIV status, TAM-TB assay format and performance appears closely compatible with the optimal TPP accuracy criteria defined by WHO for a non-sputum confirmatory TB test.

## INTRODUCTION

Early TB diagnosis will be required to reach the goal set by the WHO END TB strategy to reduce TB incidence rate by 80% in 2030 [1, 2]. Currently endorsed TB diagnostics tests encompassing smear microscopy, microbiological cultures and molecular methods all rely on sputum specimen. Diagnostic accuracy of sputum-based tests is often challenged by the paucibacillary forms of HIV-related as well as childhood TB in particular [3, 4]. The development of non-sputum-based assays relying on blood, stool or urine is urged to increase detection rate in children and people living with HIV for whom mortality remain particularly high [5].

The urine-based LAM-assay has been endorsed by WHO for TB diagnosis in HIV-infected individuals as sensitivity increases significantly in patients with lower CD4 cell counts [1]. Blood transcriptional markers and cell-free DNA in urine constitute promising additional candidates for non-sputum triage or confirmatory TB tests [6] [7]. Tuberculin skin test (TST) and interferon-gamma release assays cannot differentiate latent infection from active disease and a decreased sensitivity has been reported in children living with HIV [8, 9]. In contrast, we and others demonstrated that T cell activation markers (TAM), notably CD27, but also CD38, Ki-67, CD153, CD161 and HLA-DR, can distinguish active from latent TB, monitor patient response to treatment and retrospectively diagnose TB in children with unprecedented accuracy [10-13]. TAM-based assays depict phenotypically *Mtb*-specific CD4^+^ T cells circulating in the periphery by flow cytometry [12]. Previous studies were however mostly conducted retrospectively relying on small sample sets of cryopreserved peripheral blood mononuclear cells (PBMC) or whole-blood samples fixed and cryopreserved post-stimulation. In addition, these studies would often use as comparator healthy controls or asymptomatic but possibly latently infected individuals [10-20]. Diagnostic accuracy studies should rather be conducted in real time and only on patients showing signs of TB (presumptive TB patients) including people living with HIV. Furthermore, the index test assessment should be conducted within laboratory settings of intended use with a limited turnaround time to minimize treatment delays [21]. Until miniaturized and dedicated portable flow cytometer devices become available, TAM-based assays will be used at district hospital levels where basic flow cytometers have been installed for CD4 T cell count purposes. In summary, the implementation of TAM-based assays beyond research settings calls for a simplified protocol fulfilling the following conditions: i) being compatible with the most basic flow cytometry apparatus; ii) excluding the need of PBMC isolation; and iii) requiring a minimal amount of blood to be compatible with TB diagnosis in children.

In this study, we aimed to implement a simplified version of a TAM-based assay, assessed in real time from a single millilitre of fresh blood to deliver results within 24h. Other investigations suggested that CD38 biomarker may perform at least equally if not better than CD27 to identify diseased individuals [11, 16]. In that context, we also aimed to compare the performance of CD27 and CD38 to diagnose active TB. Following the STARD guidelines [22], we aimed to report the diagnostic performance of this assay during clinical care of a large prospective cohort of adult TB patients and controls conveniently recruited at a district-level diagnostic centre in a resource-limited setting.

## METHODS

### Study design and participants

A case-control study was conducted between September 2018 and March 2020, nested within an ongoing prospective cohort study recruiting adult (≥18 years) presumptive TB patients in the Temeke District of Dar-es-Salaam, Tanzania (TB DAR). To meet clinical team capacity constraints, the recruitment of GeneXpert^®^ negative controls was only conducted between May and December 2019. The study protocol was approved by the institutional review board of the Ifakara Health Institute, the Medical Research Coordinating Committee of the National Institute of Medical Research in Tanzania and the Ethics Committee Northwest and Central Switzerland. Study participants were recruited from National Tuberculosis and Leprosy Program (NTLP) clinic at Temeke Regional Referral Hospital, following national TB screening guidelines [23]. All participants provided a signed informed consent to collect clinical data, sputum and blood. GeneXpert^®^ MTB/RIF results from NTLP were known at time of recruitment, yet another sputum sample was collected prior initiation of treatment for culture assessment to provide a composite microbiological reference standard (see microbiological procedures). All study participants received further follow up phone interviews and clinic consultations at 2 and 5 months after enrolment and TB treatment initiation.

### Microbiological procedures

Microbiological confirmation of *Mtb* presence in patients’ sputum by GeneXpert^®^ or culture was used as the reference standard. GeneXpert^®^ results including cycle threshold (Ct) values were obtained directly from NTLP. For culture testing, all study participants provided an early morning sputum specimen prior initiation of treatment processed at the TB laboratory of the Ifakara health Institute (IHI). Sputum was treated with cetylpyridinium chloride at 25^°^C for 4-7 days to increase culture recovery as reported previously [24], before decontamination with 1% NaOH final and inoculation on Lowenstein–Jensen media at 37^0^C. Cultures were observed weekly up to 8 weeks to attest the absence of growth. A composite result including a negative GeneXpert^®^ result and a negative sputum culture result was used to rule out active TB.

### TAM-TB assay

One millilitre of fresh blood was mixed with 16 volumes of red blood cell lysis buffer (Biolegend, #420301) diluted with water (Sigma, #W3500). Nucleated cells were recovered by centrifugation and split across four sterile snap cap tubes (Corning, #352054) in 200 µl of complete medium (RPMI-1640 (Sigma, #R8758), 10% heat-inactivated FBS (Biowest, #S1810)). Cells were respectively stimulated with 1 µl of DMSO (Sigma-Aldrich, #D2650; negative control), or 1 µl of peptide pool covering the sequences of ESAT-6, CFP-10 and TB10.4 (0.25 mg/ml in DMSO; BEI Resources, NIAID, NIH, NR-50711, NR-50712 and NR-34826) in duplicates, or 2.5 µl of *Mtb* H37Rv whole cell lysate (1mg/ml in PBS; BEI Resources, NIAID, NIH: NR-14822, 1 mg/ml in PBS). After 2h, 50 µl of complete medium containing brefeldin A (Biolegend, #420601) were added before overnight stimulation. Subsequently, staining with anti-CD38-PE (clone HIT2) or anti-CD27-PE (clone O323) or an IgG1κ isotype control-PE (clone MOPC-21) was performed for 20 min. Cells were fixed and permeabilized following manufacturer’s recommendations (Biolegend buffers, #420801 & #421002) before 30 min incubation with anti-CD3-FITC (clone OKT3), anti-CD4-FITC (clone RPA-T4), anti-CD8a-APC (clone HIT8a), anti-INF-γ-PerCP (clone 4S.B3) and anti-TNF-α-PerCP (clone Mab11). Compensation particles (BD Biosciences, #552843) were generated with the above-listed antibodies. Samples were acquired on a BD FACSCalibur flow cytometer and data analysed using FlowJo_V10. The gating strategy used to determine the CD27 or CD38 phenotype of cytokine-producing T cells is provided as supplementary material. Responses were considered significant when the frequency of cytokine-producing cells was twice higher than the negative control and a minimum of five cells could be used for phenotyping (supplementary material). In the absence of a significant T cell response, the index test result was interpreted as TB disease absent (“Not TB”).

### Analysis

The diagnostic accuracy of the index tests (CD27 or CD38-based TAM assay) against the composite microbiological reference standard is reported following the STARD guidelines [22]. pROC and plotROC R packages were used to perform and plot receiver-operating characteristics analysis as well as 95% confidence intervals. The Youden method was used to define the optimal cut off value for assay sensitivity and specificity calculation (Supplementary material, panel B). Statistical comparison of diagnostic test accuracy respectively based on CD38 or CD27 marker was performed using DeLong’s method. Generalized Linear Models Fitting and a binomial family function (R version 4.0.3) was performed to identify potential association between HIV status, body mass index (BMI), age and sex and patients’ category stratified by GeneXpert^®^ results.

## RESULTS

### Study participants

Between September 2018 and March 2020, 479 GeneXpert^®^ positive cases and 108 GeneXpert^®^ negative controls were recruited from presumptive TB patients attending the National TB and leprosy clinic in Dar es-Salaam (Figure1). Baseline and clinical characteristics of study participants are summarized in Table1. Participants had a median age of 34 years [Interquartile range (IQR): 26-41] of which 17% (168/587) were females. Among study participants evaluated with presumptive TB, 17.7% were HIV-positive at enrolment of which 76 (97.45%) were already on antiretroviral therapy. Participants’ median BMI was significantly lower among GeneXpert^®^ positive patients (18.31 kg/m^2^, [IQR: 17-20.87] *vs* 21.46 kg/m^2^, [IQR: 19.11-23.73], p<0.001). The proportion of cigarette smokers was lower among GeneXpert^®^ positive study participants (23.2% *vs* 32.4%, p=0.02). Of the 479 TB patients, 12 (2.5 %) were previously treated against TB and one was identified as a treatment failure at the time of recruitment. Out of 108 GeneXpert^®^ negative study participants, 2 (1.85%) had positive microbiological results and were classified as TB patients. Multivariate logistic regression analysis (Supplementary material) adjusted for age, sex, HIV and BMI revealed that low BMI (≤ 18.5) was associated with 14% increase odds of GeneXpert^®^ positivity (Odds ratio [OR]: 1.14; 95% CI: 1-08-1.21, p<0.001). In addition, 35-44 years and over 45 years age groups were found to be respectively associated with 11% and 17% decrease odds of GeneXpert^®^ positivity compared with participants younger than 35 years of age (OR: 0.89; 95% CI: 0.81-0.98, p=0.02 and OR: 0.83; 95% CI: 0.75-0.92, p=0.001).

**Figure 1:**
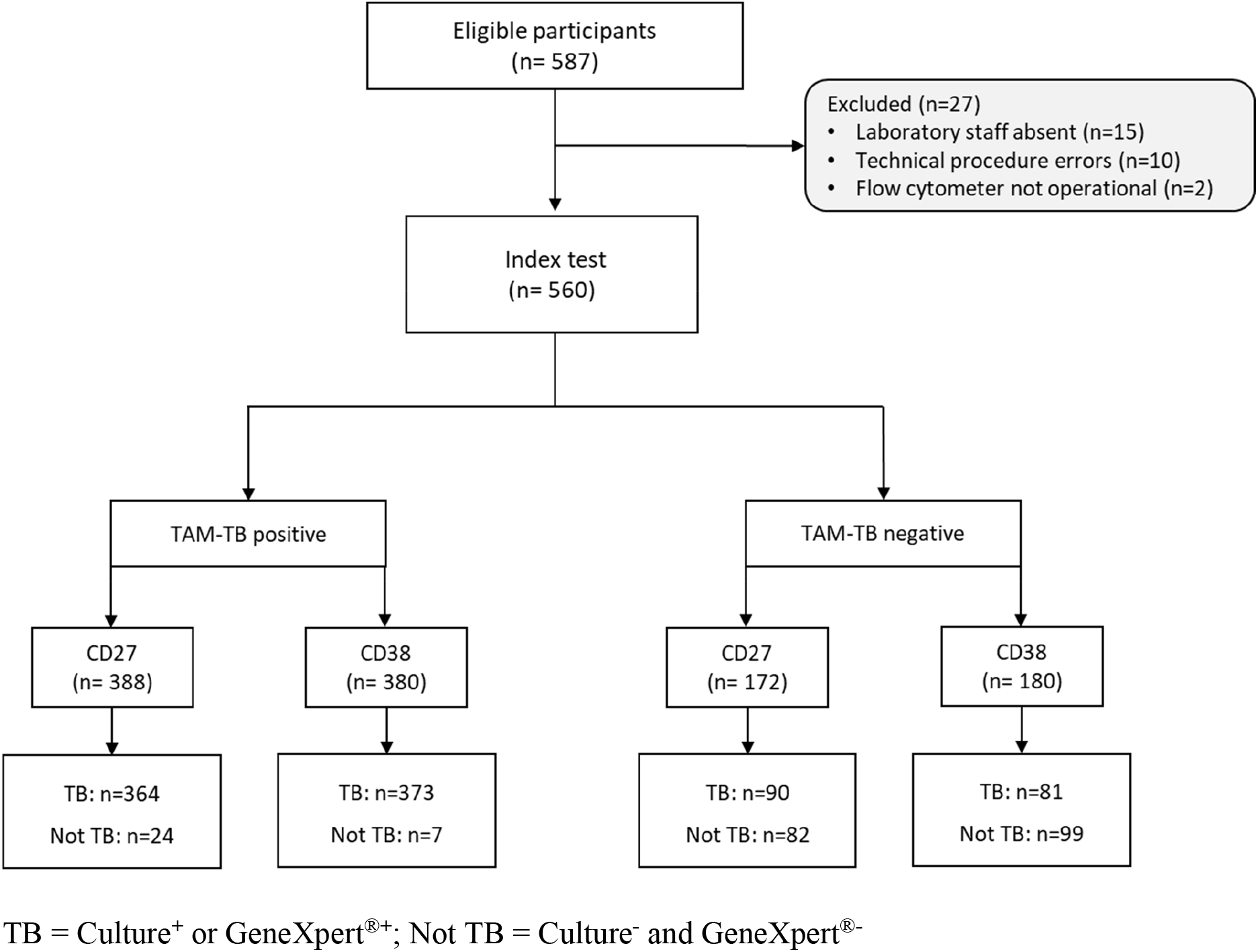
STARD flow chart showing the composition of the initial and the final study populations.

**Table 1:** Demographics and clinical characteristics of study participants stratified by GeneXpert^®^

| Characteristics | All participants<br>(n=587) | Cases | Controls |
| --- | --- | --- | --- |
|  |  | GeneXpert® positive<br>(n=479) | GeneXpert® negative<br>(n=108) |
| **Age in years, Median (IQR)** |  |  |  |
|  | 34 (26-41) | 33 (25-40) | 39 (31-45) |
| **Age group in years, n (%)** |  |  |  |
| 18-24 | 113 (19.3) | 105 (21.9) | 8 (7.4) |
| 25-34 | 200 (34.1) | 174 (36.3) | 26 (24.1) |
| 35-44 | 170 (28.9) | 129 (26.9) | 41 (37.9) |
| ≥45 | 104 (17.7) | 71 (14.8) | 33 (30.6) |
| Female, n (%) | 168 (28.6) | 133 (27.7) | 35 (32.4) |
| **BMI, kg/m<sup>2</sup>, median (IQR)** | 18.66 (17.00-20.87) | 18.31 (16.80-20.05) | 21.46 (19.11-23.73) |
| **Smoker, n (%)** | 146 (24.9) | 111 (23.2) | 35 (32.4) |
| **HIV<sup>+</sup>, n (%)** | 104 (17.7) | 78 (16.3) | 26 (24.1) |
| On ART, n (%) | na | 76 (97.4) | nd |
| **Symptoms<sup>1</sup>, n (%)** |  |  |  |
| Cough | 576 (98.1) | 473 (98.7) | 103 (93.4) |
| Fever | 464 (79) | 398 (83.1) | 66 (61.1) |
| Night sweat | 327 (55.7) | 298 (62.2) | 29 (26.9) |
| Significant weight loss | 424 (72.2) | 381 (79.5) | 43 (39.8) |
| **TB patient category, n (%)** |  |  |  |
| New | 468 (97.3) | 466 (97.3) | 2* (1.8) |
| Relapse | 12 (2.5) | 12 (2.5) | - |
| Treatment after default | 1 (0.2) | 1 (0.02) | - |
| **Full blood counts<sup>1</sup> (10<sup>9</sup> cells/L)** |  |  |  |
| White Blood cells, median (IQR) | 7.23 (5.78-9.04) | 7.55 (6.12-9.46) | 5.95 (4.79-7.14) |
| Platelets, median (IQR) | 323 (243-417) | 344 (262-430) | 251 (208-305) |
| Red blood cells, mean (±SD) | 4.61 (0.85) | 4.61 (0.87) | 4.59 (0.78) |
| **Culture results, n (%)** |  |  |  |
| Scanty | 29 (4.9) | 27 (5.6) | 2* (1.8) |
| 1<sup>+</sup> | 231 (39.6) | 231 (48.2) | - |
| 2<sup>+</sup> | 136 (23.2) | 136 (28.4) | - |
| 3<sup>+</sup> | 30 (5.1) | 30 (6.3) | - |
| Contaminated | 2 (0.34) | 2 (0.41) | - |
| Negative | 145 (24.7) | 41 (8.6) | 104 (96.3) |
| Missing | 14 (2.4) | 12 (2.5) | 2 (1.8) |
| **GeneXpert®<sup>1</sup>, n (%)** |  |  |  |
| MTB detected | 475 (99.2) | 475 (99.2) | - |
| MTB not detected | 108 (100) |  | 108 (100) |
| MTB detected/Rif resistance | 4 (0.8) | 4 (0.8) | - |

BMI-Body mass index, Culture results; Scanty: < 20 colonies, 1<sup>+</sup>: 20 to 200 colonies, 2<sup>+</sup>: > 200 discrete colonies, 3<sup>+</sup>: > 200 confluent colonies. \* Recruited as controls but included as cases, MTB: *M. tuberculosis*, RIF: Rifampicin, ART: Antiretroviral therapy; <sup>1</sup>Symptoms/condition at enrolment, na: not applicable, nd: no data.

### TAM-TB diagnostic performance

The diagnostic performance of CD38-versus CD27-based TAM assays was evaluated side by side across 560 presumptive TB patients for whom a TB diagnosis was confirmed or not by GeneXpert^®^ or culture (STARD flow chart, figure1). The use of an isotype control antibody in the negative control allowed us to determine both positivity threshold for the expression of the investigated biomarkers and for cytokine detection (gating strategy provided as supplementary material, panel B). The TAM-based assay value here reflects a ratio between the frequency of *Mtb* peptide pool-specific CD4 T cells expressing the biomarker of interest and those that did not. Compared to our previous report that used a median intensity ratio of biomarker expression [10], this approach increased significantly the performance of the CD38-based TAM assay and revealed that a minimum of five cytokine-producing T cells was required to optimally assess TB status in this cohort (Supplementary material, panel C). In addition, receiver operating characteristic analysis revealed a significantly superior diagnostic accuracy of the CD38-based index test (Area under the curve, AUC=0.87, 95%CI 0.84-0.91) compared to the CD27-based index test (AUC=0.81, 95%CI 0.77-0.85) (p=0.003) (Figure 2). The CD38-based assay was able to rule out TB in 99 of 106 non-TB patients and properly diagnosed TB in 373 of 454 TB patients resulting in an assay sensitivity and specificity of 82.2% and 93.4% respectively, at an assay cut-off value of 0.74 (Figure 2). HIV infection status did not significantly impact the performance of the index test for any of the two markers investigated. Furthermore a multivariable logistic regression indicated that age, sex, BMI and smoking status did not influence specificity and/or sensitivity of the index test.

**Figure 2:**
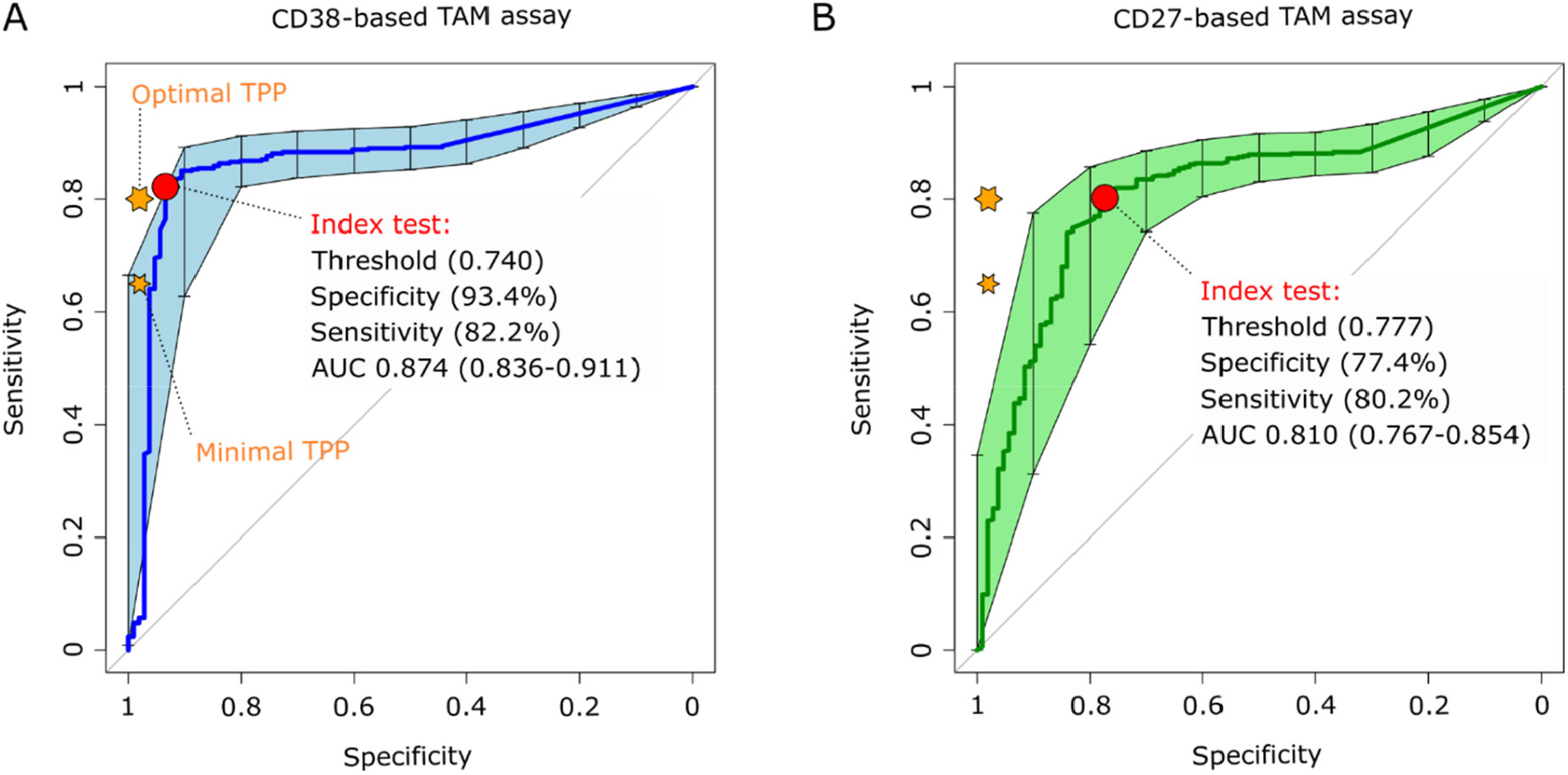
Receiver operating characteristic (ROC) curves of CD38-based (left plot) and CD27-based index test. The orange stars indicate the target product profile (TPP) minimal (80/98) and optimal (65/98) sensitivity/specificity values defined by WHO for non-sputum confirmatory TB diagnostic tests. The red circles indicate the index test performance and threshold values yielding the best specificity for a test sensitivity above 80%, the minimal TPP requirement. AUC, area under the cure and 95% confidence intervals within brackets.

We then sought to investigate the patients’ characteristics that may have driven some of the false positive and negative results of the CD38-based index test (Table 2). Among 7 presumptive TB patients with a false positive CD38-based TAM assay, one was lost on follow-up and one was found to have received TB treatment based on subsequent GeneXpert^®^ positive testing between enrolment and our follow-up visit increasing retrospectively the index test specificity from 93.4 to 94.3%. False negative TB patients did not show a significantly decreased BMI or HIV infection rate that may have sustained a hampered immune reactivity in vitro. Noteworthy, 6.2% of index test true positive TB patients had negative culture results compared to 18.5% of false negatives. This also translated into a slight increase in the averaged GeneXpert^®^ cycle threshold of false negative compared to true positive TB patients.

**Table 2:** Patients and assay characteristics at enrolment stratified by CD38-based index test results

| Characteristics | CD38-based index test |  |  |  |
| --- | --- | --- | --- | --- |
|  | False positive<br>(n=7) | False negative<br>(n=81) | True positive<br>(n=373) | True negative<br>(n=99) |
| **Age in years, Median (IQR)** | 32<br>(28.5-43.5) | 36<br>(29-41) | 32<br>(25-39) | 39<br>(32-45) |
| **Female, n (%)** | 2 (28.6) | 13 (16) | 112 (30.1) | 33 (33.3) |
| **BMI, kg/m2, median (IQR)** | 22.79<br>(19.17-24.06) | 18.67<br>(17.27-20.28) | 18.14<br>(16.71-19.95) | 21.34<br>(19.15-23.67) |
| **Smoker, n (%)** | 1 (14.3) | 25 (30.9) | 84 (22.5) | 32 (32.3) |
| **HIV<sup>+</sup>, n (%)** | 2 (28.6) | 11 (13.5) | 64 (17.2) | 24 (24.2) |
| **On ART, n (%)** | - | 11 (100) | 62 (96.9) | - |
| **Culture results, n (%)** |  |  |  |  |
| Negative | 7 (100) | 15 (18.5) | 23 (6.2) | 97 (98) |
| Scanty | - | 3 (3.7) | 25 (6.7) | - |
| 1<sup>+</sup> | - | 36 (44.4) | 176 (47.2) | - |
| 2<sup>+</sup> | - | 20 (24.7) | 112 (30) | - |
| 3<sup>+</sup> | - | 4 (4.9) | 26 (6.9) | - |
| Contaminated | - | 2 (2.5) | 11(3) | - |
| Missing | - | 1 (1.2) | - | 2 (2) |
| **GeneXpert®** |  |  |  |  |
| Positive, n (%) | - | 80 (98.8) | 372 (99.7) | - |
| Cycle threshold (IQR) | - | 19.15<br>(16.52-23.95) | 18.20<br>(15.40-21.40) | - |
| Negative, n (%) | 7 (100) | 1(1.2) | 1(0.3) | 99 (100) |
| **Month 5 follow-up, n (%)** |  |  |  |  |
| Improved or recovered | 5 (72.4) | 59 (72.8) | 284 (76.1) | 62 (62.6) |
| Interim TB treatment | 1 (14.3) | - | - | - |
| Lost | 1 (14.3) | 22 (27.2) | 89 (26.9) | 35 (35.4) |
| **Index test negativity** |  |  |  |  |
| Cytokine resp. < cut-off | na | 58 (71.6) | na | 77 (77.8) |
| Phenotypic resp. < cut-off | na | 23 (28.4) | na | 22 (22.2) |

BMI-Body mass index. Scanty: < 20 colonies, 1<sup>+</sup>: 20 to 200 colonies, 2<sup>+</sup>: > 200 discrete colonies, 3<sup>+</sup>: > 200 confluent colonies. MTB: *M. tuberculosis*, RIF: Rifampicin, ART: Antiretroviral therapy; <sup>1</sup>Symptoms/condition at enrolment, na: not applicable, nd: no data.

## DISCUSSION

Our study has strengths and limitations. First, the study team was responding to the delivery of clinical samples in the context of clinical care in a completely unforeseen manner, processing from none up to 6 samples daily. This is in contrast to most if not all previous studies that cumulated whole blood stimulated samples or PBMCs which were processed later in batches within optimal experimental conditions [10-20]. The real time assessment reported in this study demonstrates the feasible implementation of a TAM assay as a routine diagnostic tool at district hospital level hosting the most basic 2-lasers flow cytometer apparatus. With a turn-around time of 24h, the implementation of such an assay has the potential to dramatically fasten decision to initiate TB treatment. Future studies including childhood and extra-pulmonary presumptive TB patients should now be pursued to ascertain its potential to indicate TB infection in these specific groups of patients. The main limitation of this study lies on the convenience of the sampling, which even though prospective, led to an unbalanced representation of presumptive TB patients from which TB had been excluded and that may have biased our interpretation of the assay specificity. In addition, and although sputum culture results were pending, the study team was not blinded from GeneXpert^®^ results while performing and analysing the index test, and this is considered to potentially overestimate the interpretation of the assay sensitivity [21]. Nonetheless, this study relies on a substantial and unprecedented sample size and format, based only on symptomatic presumptive TB patients. In addition, the index test only required one millilitre of blood that can be obtained from finger or heel pricks which supports a realistic potential of TAM-based assay to deliver a point of care diagnostic test for childhood TB within 24h.

We observed that the CD38-based assay performed significantly better than CD27 to differentiate active TB from non-TB patients (Figure 2). The poor performance of CD27 contrasts our previous observation [10], where a CD27-based assay could achieve 96.8% specificity and 83.3% sensitivity in a cohort of children presumptive TB patients. This may arise from differential immune maturation processes in children and adults, but also from the inclusion of TNF-α producing CD4 T cells in the phenotypic assessment of the TAM assay. The poor performance of the CD27-based assay in adults may also rely on the observation reported by Halliday *et al*. that it cannot distinguish recently from remotely acquired *Mtb* infection [25]. As such, CD27 may be more influenced by infection than disease explaining why it seemed to perform better in young children, where infection is less common among controls. Although significantly superior to CD27, the specificity of the CD38-based assay was still hampered by clear signals of recent exposure in seven of the presumptive TB patients, from which *Mtb* could not be characterised by culture or molecular detection. Interestingly, upon invitation for a follow-up visit of index test positive patients, one of them had initiated TB treatment based on further interim GeneXpert^®^ testing at NTLP (Table 2). To fully exclude that the remaining false positive presumptive TB patients may not represent cases of incipient TB, careful clinical monitoring would be required for a minimum of two years [26]. Yet, our data suggest that such patients would likely benefit from TB chemotherapy. In return, the CD38-based TAM assay diagnosed TB for two GeneXpert^®^ negative controls for which TB status was only later assigned by culture. Both patients were males, had a history of direct contact with an index case in their neighbourhood, were HIV negative and displayed symptoms suggestive of TB (cough and/or fever, haemoptysis and night sweats).

By depleting CD4 T cells, HIV infection may logically reduce the performance of blood-based immune assays [8, 27]. When we compared the TAM assay performances between HIV-infected and HIV-negative groups, we could not detect any significant difference between the two groups. The relatively low rate of HIV co-infection in our cohort and the fact that a large majority of HIV-positive study participants were already on anti-retro viral treatment at the time of enrolment may have hampered this assessment. Nonetheless, our findings supports the conclusion by Wilkinson *et al*, [28] that T cell activation markers can distinguish active and latent TB infection irrespective of HIV status. Of note, the observed performance of the CD38-based TAM assay reported here bypassed the minimal 80% sensitivity and closely reached the 98% specificity specified for the optimal Target Product Profile (TPP) prioritized by WHO for non-sputum-based TB diagnostic tests [5]. These results should further motivate the assessment of CD38-based assays in diagnostic accuracy studies including children as well as extra-pulmonary presumptive TB patients.

At this stage, the prospective implementation of TAM-based assays relies on the availability of a flow cytometry platform and availability of trained laboratory technicians to run the assay and analyse its results. Together with the development of dedicated kits being pursued by Beckman Coulter and LMU Munich (Communicated at StopTB annual meeting 2020), the previously reported miniaturisation of flow cytometry devices [29, 30] and the development of tools to automate high-throughput analysis of clinical flow cytometry data [31], the development of a TAM assay for TB diagnosis seems reasonably at reach. Nonetheless, several other non-sputum-based diagnostic tools including host RNA or protein signatures [32, 33], as well as cell-free DNA detection in urine are also promising [34]. Concerted and side-by-side evaluation of these different approaches across multiple sites should be performed to urgently prioritize the development of non-sputum TB diagnostic tools.

## CONCLUSIONS

The presented data confirms the feasibility and potential of implementing the TAM-TB assay as a point of care test for TB diagnostics from a single millilitre of blood. The test will not inform on the drug sensitivity profiles. Yet, with a 24h turnaround time the TAM-TB assay constitutes an excellent candidate for early TB detection in specific high-risk groups with extra-pulmonary or paucibacillary forms of TB, such as HIV patients and children.

## Data Availability

TB DAR cohort study constitutes an observational study that did not require submission to TMDA but an ethical clearance was obtained as detailed above. The data used in this manuscript are not publicly available due to the Tanzania national policy on data sharing. Data will be made available upon request where concerned parties will sign a data transfer agreement approved by the Medical Research Coordinating Committee.

## List of abbreviations

ART: Antiretroviral therapy
AUC: Area under the curve
BMI: body mass index
Ct: cycle threshold
NTLP: National Tuberculosis and Leprosy Program
PBMC: Peripheral Blood Mononuclear Cells
RIF: Rifampicin
TAM: T cell activation marker
TB: tuberculosis
TPP: target product profiles

## Acknowledgement

We thank all study participants who willingly participated in this study. We are grateful to all the staff and administration of the Temeke Regional Referral Hospital especially the NTLP clinic for their diligent work and support during recruitment. We are also grateful to our study team at the Temeke clinic and Ifakara health institute Bagamoyo laboratory for recruiting patients, sampling, and collecting data.

## Data availability/accessibility/where study protocol can be accessed

TB DAR cohort study constitutes an observational study that did not require submission to TMDA but an ethical clearance was obtained as detailed in the methods section. The data used in this manuscript are not publicly available due to the Tanzania national policy on data sharing. Data will be made available upon request where concerned parties will sign a data transfer agreement approved by the Medical Research Coordinating Committee.

