## Supplementary material for "Case-control, diagnostic accuracy study of a non-sputum CD38-based TAM-TB test from a single milliliter of blood"

**Supplementary material.** A) TAM-based assay procedure overview from blood collection to delivery of results within 24h. B) Representative gating strategy of the TAM-based assay adapted for a 2-laser flow cytometer FACSCalibur apparatus. First, a gate based on Forward-(FSC) and Side-Scattered (SSC) signals is drawn to isolate lymphocytes from other blood cell types. Second, a CD4 T cell gate is obtained by tearing apart non T cells in the y-axis from CD8 T cells in the x-axis. This gating strategy is applied to compare cytokine production and expression of CD27 or CD38 by CD4 T cells in samples stimulated or not with a *Mycobacterium tuberculosis* peptide pool. The index test result is obtained by dividing the frequency of CD4 T cells producing IFN-γ or TNF-α that express the investigated biomarker (Quadrant 2, Q2) divided by the frequency of those that did not express the biomarker (Q3). C) The area under the Receiving Operating Characteristics curve (AUC) was extracted to determine that compared to a Median Fluorescence Intensity ratio approach used previously for CD27 [10], the Q2/Q3 ratio and a minimum of five cytokine-producing CD4 T cells were required to optimally diagnose TB in this cohort of presumptive TB patients. D) Risk factors for GeneXpert® positivity and GeneXpert® negativity: Body mass index (BMI); Human immunodeficiency virus (HIV); Unadjusted Odds ratios (ORs); Adjusted Odds ratio (aORs); Logistic regression models (univariate and multivariable logistic regression) was performed to determine association between binary outcome (GeneXpert® positive and GeneXpert® negative) and age in years, sex, body mass index, smoking and HIV status.

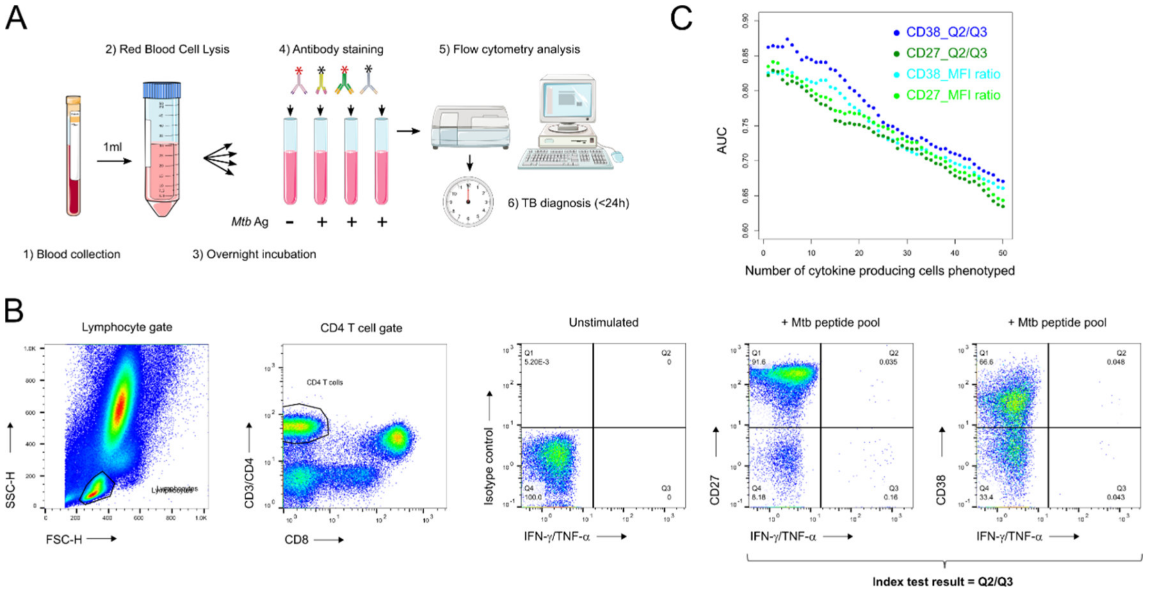


D

| Characteristics | GeneXpert® status | | Unadjusted | | Adjusted | |
|---|---|---|---|---|---|---|
| | positive, n(%) | negative, n (%) | OR (95%CI) | p-value | aOR (95%CI) | p value |
| **Age group (years)** | | | | | | |
| 18-24 | 105 (21.9) | 8 (7.4) | Ref | | Ref | |
| 25-44 | 174 (36.3) | 26 (24.1) | 0.94(0.86-1.03) | 0.18 | 0.98 (0.90-1.08) | 0.77 |
| 35-44 | 129 (26.9) | 41 (37.9) | 0.84(0.77-0.92) | 0.001 | 0.89(0.81-0.98) | 0.02 |
| >45 | 71 (14.8) | 33 (30.6) | 0.78(0.71-0.86) | <0.001 | 0.83(0.75-0.92) | 0.001 |
| **Sex** | | | | | | |
| Female | 133 (27.7) | 35 (32.4) | Ref | | Ref | |
| Male | 346 (72.3) | 73 (67.6) | 1.04 (0.96-1.11) | 0.33 | 1.05 (0.98-1.13) | 0.16 |
| **BMI category (kg/m²)** | | | | | | |
| BMI≥18.5 | 220 (45.9) | 87 (80.1) | Ref | | Ref | |
| BMI<18.5 | 259 (54.1) | 21 (19.9) | 1.23(1.16-1.31) | <0.001 | 1.14(1.08-1.21) | <0.001 |
| **Smoking status** | | | | | | |
| no | 368(76.8) | 73(67.6) | Ref | | Ref | |
| yes | 111 (23.2) | 35 (32.4) | 0.93(0.86-0.99) | 0.045 | 0.94(0.87-1.02) | 0.12 |
| **HIV status** | | | | | | |
| Negative | 401(83.7) | 82(75.9) | Ref | | Ref | |
| Positive | 78 (16.3) | 26 (24.1) | 0.92(0.85-1.00) | 0.055 | 0.95(0.88-1.03) | 0.24 |
